# Dementia Risk and Machine Learning-Derived Brain Age Index from Sleep Electroencephalography: A Pooled Cohort Analysis of Over 7,000 Individuals Across Five Community Cohorts

**DOI:** 10.1101/2025.09.21.25336255

**Authors:** Haoqi Sun, Sasha Milton, Yi Fang, Hash Brown Taha, Shreya Shiju, Robert J. Thomas, Wolfgang Ganglberger, Matthew P. Pase, Timothy Hughes, Shaun Purcell, Susan Redline, Katie L. Stone, Kristine Yaffe, M. Brandon Westover, Yue Leng

## Abstract

**Importance:** Sleep electroencephalographic (EEG) microstructures are closely related to cognition and undergo age-dependent changes. However, their multidimensional nature makes them challenging to interpret using conventional approaches. Machine learning-computed EEG brain age index (BAI) represents the difference between the sleep EEG-based brain age and chronological age, quantifying deviations in sleep EEG microstructures from normative patterns.

**Objective:** To determine the association between sleep BAI and incident dementia in community-dwelling populations.

**Design:** Five individual cohorts and random-effects meta-analysis.

**Setting:** This study pooled data from five community-based, methodologically consistent, longitudinal cohorts: MESA, ARIC, FHS-OS, MrOS, and SOF. We used Fine-Gray models to assess the association between BAI and incident dementia within each cohort, accounting for death as a competing risk. Cohort-specific estimates were then pooled using random-effects meta-analyses.

**Participants:** 7,071 participants (MESA 54-94 years old, ARIC 52-75, FHS-OS 40-81, MrOS 67-96, SOF 79-93) without dementia at the time of polysomnography were included.

**Exposure:** The sleep EEG-based BAI was computed using interpretable machine learning, incorporating 13 age-dependent features extracted from central EEG channels in overnight, home-based sleep polysomnography.

**Main Outcomes and Measures:** Incident dementia or probable dementia was determined in each cohort, with death as a competing risk.

**Results:** Across the five cohorts, dementia incidence ranged from 6.6% to 34.3% over a median follow-up of 3.5 to 17.0 years. Across cohorts, each 10-year increase in BAI was associated with a 39% increased risk of incident dementia (hazard ratio: 1.39 [95% confidence interval=1.21–1.59], p<0.001) after adjustment for age, sex, race, education, body mass index, current smoking, sleep medications, and physical activity level. The top feature underlying BAI was waveform kurtosis in N2 with a negative association with incident dementia (p<0.001). The associations remained after additional adjustment for multiple comorbidities, APOE e4 status, and apnea-hypopnea index, and were consistent across sex and age groups.

**Conclusions and Relevance:** A higher sleep EEG-based BAI was associated with a higher risk of incident dementia across five community-based longitudinal cohorts. Future studies are warranted to evaluate the predictive value of BAI as a non-invasive digital biomarker for the early detection of dementia in community settings.

## Introduction

Dementia is an important public brain health concern with profound impacts on individuals and society, including in low- and middle-income countries^1^. Meanwhile, sleep is an important component of brain health^2^. Both overly short and long sleep durations, as well as sleep disorders, are associated with incident dementia^3–5^, including Alzheimer’s disease (AD)^6^. However, the macro-level sleep architecture has shown inconsistent associations with cognitive impairment and incident dementia^7,8^. These broad sleep metrics do not fully capture the complex and multidimensional nature of sleep physiology. In contrast, the microstructure of sleep electroencephalography (EEG) directly reflects the neural processes with explicit functional implications^2^, presenting an opportunity to develop digital prodromal markers for the early detection of dementia and provide more nuanced insights into cognitive aging^9–11^.

Prior studies have shown that cognitive impairment is associated with multiple sleep EEG patterns, including spectral power^12–14^, sleep depth^15,16^, and spindle / slow oscillation (SO) / coupling^11,12,17^. Despite the promising insights provided by sleep EEG, the vast amounts of EEG patterns make it challenging to summarize and interpret. One innovative approach is to quantify deviations from normal aging patterns. For instance, the frequency of the alpha posterior dominant rhythm peaks at 10-11 Hz around age 30, and gradually decreases to 8-9 Hz by age 80^18^, and spindle density decreases with age^10,19^. While sleep EEG microstructural changes typically follow a predictable pattern with normal aging, some individuals may experience accelerated deterioration in these patterns. Sleep EEG microstructure that matches an age older than an individual’s chronological age may indicate accelerated brain aging and risk for worse cognitive outcomes.

To integrate these complex patterns, we developed a sleep EEG-based brain age using a novel interpretable machine learning approach, integrating multiple age-dependent EEG microstructures into an age-like number^20^. The difference between brain aging and chronological age is termed the brain age index (BAI). While elevated sleep EEG-based BAI has been associated with dementia in a clinical-based cross-sectional study^21^, this prior study has important limitations, including the use of subjects referred for a clinical assessment in a sleep clinic, and thus not representative of the general population, while the cross-sectional designs limit the ability to assess temporality. It remains unclear if BAI is associated with incident dementia in community-dwelling populations.

Here, we computed the BAI from sleep EEG microstructures^20^ and examined its association with incident dementia across five community-dwelling longitudinal cohorts using individual participant data and random-effects meta-analysis. We examined whether the association between the BAI and dementia risk differed by age and sex, and if the association was influenced by key dementia risk variables, including APOE e4 carrier status and comorbidities. Additionally, we examined the association between individual EEG features underlying the BAI model and dementia risk to interpret their relative contributions and gain insight into the underlying neurophysiology.

## Methods

### Study Design

The five community-based prospective cohorts included the Multi-Ethnic Study of Atherosclerosis (MESA)^22^, the Atherosclerosis Risk in Communities (ARIC) study^23^, the Framingham Heart Study Offspring cohort (FHS-OS, i.e., FHS Gen 2)^24^, the Osteoporotic Fractures in Men (MrOS) Study^25,26^, and the Study of Osteoporotic Fractures (SOF)^27^. Details on each cohort are in the Supplementary Methods. For all cohorts, sleep data were collected using a similar protocol overseen by the same sleep reading center (directed by SR), where data were processed and scored similarly. As in Figure 1, the inclusion criteria were: (1) availability of overnight sleep PSG; and (2) availability of outcomes, including time-to-event data and event type (dementia, death, censored); the exclusion criteria were: (1) missing BAI due to absence of spindles or excessive artifacts (defined as epochs with amplitude >500uV or standard deviation (SD) <0.1uV); (2) missing covariates (see below); and (3) prevalent dementia at the time of the sleep study.

**Figure 1.**
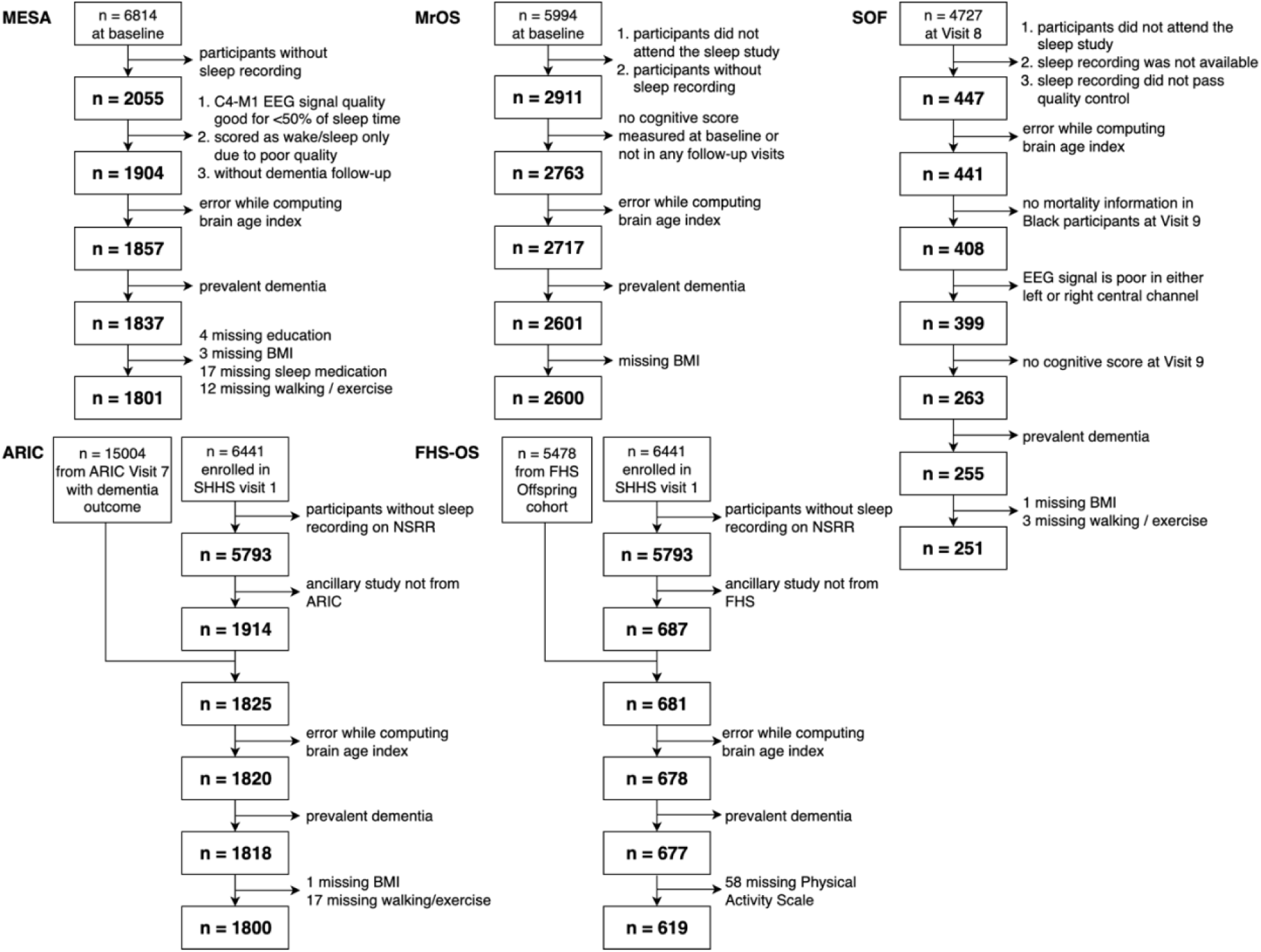
Flowchart of the included studies. MESA – Multi-Ethnic Study of Atherosclerosis. MrOS – Osteoporotic Fractures in Men. SOF – Study of Osteoporotic Fractures. ARIC – Atherosclerosis Risk in Communities. BMI – body mass index. EEG – electroencephalography. FHS-OS – Framingham Heart Study Offspring. NSRR – National Sleep Research Resource.

All cohort committees approved the use of the data. Informed consent was obtained in each cohort. These analyses followed reporting guidelines specified by Strengthening in Reporting of Observational Studies in Epidemiology (STROBE)^28^. The analysis was conducted between March 2024 and September 2025.

### Outcomes

The primary outcome was incident dementia, with death treated as a competing risk. Details on dementia and death ascertainment are in the Supplementary Methods. The methods used to determine dementia varied across cohorts and included both clinical adjudication and cognitive score-based approaches at periodic follow-ups. Briefly, in ARIC, FHS-OS, and SOF, dementia adjudication was reached through expert panels with serial neuropsychological tests and informant interviews, involving the Diagnostic and Statistical Manual (DSM)^29^ and other criteria. In MESA, dementia was based on hospitalized ICD-based all-cause dementia^30^ or a decrease in Cognitive Abilities Screening Instrument (CASI, a global cognitive score)^31^ ≥ 1.5 standard deviations across participants from Exam 5 (close to sleep study) to Exam 6. In MrOS, probable dementia was ascertained based on self-report of physician diagnosis, use of dementia medication, or a decrease in Modified Mini-Mental State (3MS)^32^ scores ≥ 1.5 standard deviations across participants from baseline to any follow-up visit.

Death was ascertained using death certificates. Participants who experienced neither dementia nor death were censored at the date of last contact. Time-to-event was defined as the number of days from the sleep study to the first occurrence of dementia, death, or censoring.

### Sleep EEG-based Brain Age Index

Participants underwent unattended in-home overnight PSG using the Compumedics System (Abbotsford, Victoria, Australia), with different models (SHHS: Series P; MrOS: Safiro; SOF: Siesta; MESA: Somte). We used the same EEG preprocessing and brain age computation as previously described^20^. In brief, we used two central EEG channels (C3-M2, C4-M1) for cohorts other than MESA and one central EEG channel (C4-M1) for MESA. EEG signals were notch-filtered at 60Hz, bandpass filtered at 0.5Hz to 35Hz, and then resampled to 200Hz. To minimize artifacts, we excluded 30-second epochs with maximum absolute amplitude >500uV or with more than 2 seconds of flat signal (SD <0.2uV).

The brain age was based on an alternative version of Sun et al as implemented in Luna^10^. We selected 13 EEG microstructure features out of the original 510 features that fulfill both high longitudinal precision and high correlation with age, based on a stepwise feature selection approach. The features were extracted from artifact-free 30-second epochs, including spindle density, spindle-SO coupling overlap, alpha (8-12Hz) band power in N1, delta (0.5-4Hz), theta (4-8Hz), alpha, sigma (12-20Hz) band power kurtosis in N2, delta band power in N3, theta band power kurtosis in N3, delta-alpha ratio and delta-theta ratio in N3, and signal waveform kurtosis in N2 and N3. These features were input into a trained brain age model, which outputs brain age as a weighted sum of the sleep EEG microstructures. The model was trained on relatively healthy participants at 18 to 80 years old from a clinical cohort without major neurological or psychiatric diseases, including neurodegenerative diseases, stroke, and epilepsy^10^. The BAI was defined as the predicted BA minus age, where a negative value indicates a younger brain age, and a positive value indicates an older brain age.

### Statistical Analysis

Within each cohort, we used the Fine-Gray subdistribution hazard model^33,34^ to assess the association between BAI and incident dementia, treating death as a competing risk. Covariates included age at the time of the sleep study, sex, race and ethnicity, education, body mass index (BMI), current smoker, use of sleep medications, and physical activity level. Table S1 provides the definitions of these covariates for each cohort, indicating that some covariates were excluded from certain cohorts and that others had varying definitions across cohorts. We fit two models for each cohort: a minimally adjusted model (adjusting for age and sex) and a fully adjusted model (additionally adjusting for race and ethnicity, education, BMI, current smoker, sleep medication, and physical activity). For each cohort, we estimated the hazard ratios (HRs) and 95% confidence intervals (CIs) for a 10-year increase in BAI. We applied Fine-Gray models to examine the associations between individual BAI features and dementia risk, with HRs calculated per 1-SD increase in each feature.

In sensitivity analyses, we adjusted for additional covariates where available: diagnoses of depression, diabetes, hypertension, heart attack, and stroke at the time of the sleep study, baseline cognition assessed as part of the cohort study, apnea-hypopnea index (AHI) (3% rule for hypopnea), and APOE e4 allele carrier. We performed stratified analyses in males and females, and younger (<70 years old) and older (≥70 years old) individuals (as 70 is the overall mean age). Cohorts without eligible participants for a given subgroup were excluded from the corresponding analyses.

Cohort-specific estimates were pooled using an inverse-variance random effects model following the Der Simonian and Laird model^35^. The Higgins I^2^ test was used to assess heterogeneity across cohorts^36^. The pooled meta-analysis mitigated the harmonization problem of the cohort-specific covariates. The analyses were performed using the *meta* package^37^ version 7.0.0 in R 4.3.2.

## Results

### Cohort Characteristics

As in Table 1, our study included a total of 7,071 participants, including 1,801 from MESA, 1,800 from ARIC, 619 from FHS-OS, 2,600 from MrOS, and 251 from SOF. Of these, 2,447 (34.6%) were females. The mean age was 70 years, ranging from 59.5 years in the FHS-OS to 82.7 years in SOF. The median time to dementia from sleep study ranged from 3.5 years in MrOS to 17.0 years in ARIC. The mean BAI ranged from -5.4 years in SOF to +0.6 years in FHS-OS. As in Figure 2, the sleep EEG-based brain ages had mean absolute errors (MAEs) ranging from 4.7 years in the FHS-OS to 6.0 years in SOF. Table S2 shows the characteristics of participants with lower BAI (≤ -3 years) and higher BAI (≥ 3 years), and their univariate comparison results.

**Figure 2.**
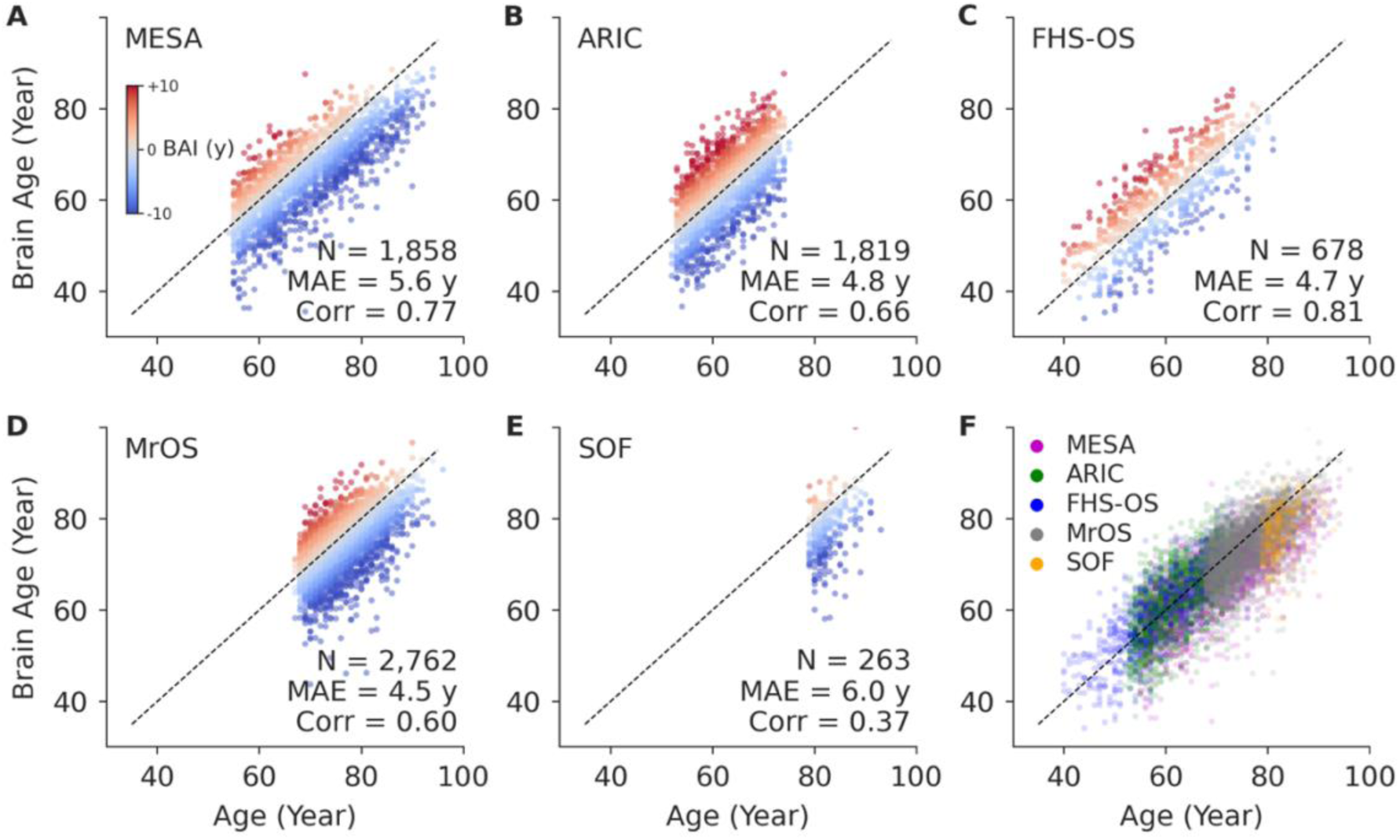
Scatter plots of chronological age vs. sleep EEG-based brain age in all cohorts. Each dot represents one participant. The diagonal line represents where age equals to brain age. The last panel show all cohorts overlaid. MAE – mean absolute error. Corr – Pearson’s correlation coefficient. MESA – Multi-Ethnic Study of Atherosclerosis. ARIC – Atherosclerosis Risk in Communities. FHS-OS – Framingham Heart Study Offspring. MrOS – Osteoporotic Fractures in Men. SOF – Study of Osteoporotic Fractures.

**Table 1.** Participant characteristics across all cohorts included in the meta-analysis.

| Name | MESA<br>(n=1,801) | ARIC<br>(n=1,800) | FHS-OS<br>(n=619) | MrOS<br>(n=2,600) | SOF<br>(n=251) |
| --- | --- | --- | --- | --- | --- |
| Age at sleep study<sup>†</sup>, year | 69.3 (9.0) | 62.5 (5.7) | 59.5 (8.9) | 76.0 (5.3) | 82.7 (2.9) |
| Female<sup>†</sup> | 955 (53.0%) | 922 (51.2%) | 319 (51.5%) | 0 (0%) | 251 (100%) |
| Race & Ethnicity<sup>†</sup> |  |  |  |  |  |
| Asian | 211 (11.7%) | 0 (0%) | 0 (0.0%) | 71 (2.7%) | 0 (0%) |
| Black | 498 (27.7%) | 0 (0.0%) | 0 (0.0%) | 75 (2.9%) | 0 (0%) |
| Hispanic | 420 (23.3%) | -- | 1 (0.2%) | -- | -- |
| White | 672 (37.3%) | 1800 (100%) | 556 (89.8%) | 2387 (91.8%) | 250 (99.6%) |
| College degree<sup>†</sup> | 725 (40.3%) | 795 (44.2%) | 387 (62.5%) | 2072 (79.7%) | 200 (79.7%) |
| BMI<sup>‡</sup>, kg/m<sup>2</sup> | 27.8 (24.7-31.7) | 28.2 (25.2-31.5) | 27.7 (25.1-31.2) | 26.8 (24.7-29.3) | 27.1 (24.6-30.6) |
| Current smoker | 125 (6.9%) | 183 (10.2%) | 72 (11.6%) | 55 (2.1%) | 3 (1.2%) |
| APOE e4 carrier among available<sup>*,+</sup> | 466 (27.2%) | -- | 81 (20.9%) | 547 (23.8%) | 11 (7.9%)<sup>§</sup> |
| Incident dementia<sup>+</sup> | 119 (6.6%) | 355 (19.7%) | 59 (9.5%) | 463 (17.8%) | 86 (34.3%) |
| Time to dementia from sleep study<sup>&</sup>, year | 4.8 (4.2-5.6) | 17.0 (14.9-19.8) | 13.1 (8.5-16.2) | 3.5 (1.3-7.1) | 4.6 (4.2-5.2) |
| BAI<sup>^</sup>, year | -4.2 (6.1) | -0.0 (6.0) | +0.6 (5.8) | -3.4 (5.6) | -5.4 (5.7) |
| AHI<sup>&</sup>, /hour | 8.6 (3.2-19.4) | 5.2 (1.9-13.0) | 4.5 (1.5-11.7) | 8.1 (3.2-16.6) | 9.6 (5.2-18.8) |
| Sleep medication use<sup>+</sup> | 200 (11.1%) | 190 (10.6%) | 50 (8.1%) | 351 (13.5%) | 48 (19.1%) |
| Hypertension<sup>+</sup> | 1015 (56.4%) | 653 (36.3%) | 204 (33.0%) | 1274 (49.0%) | 148 (59.0%) |
| Diabetes<sup>+</sup> | 345 (19.3%) | 97 (5.4%) | 35 (5.7%) | 334 (12.8%) | 26 (10.4%) |
| Myocardial infarction<sup>+</sup> | 35 (1.9%) | 112 (6.3%) | 33 (5.4%) | 432 (16.6%) | 33 (13.1%) |
| Stroke<sup>+</sup> | 25 (1.4%) | 42 (2.3%) | 7 (1.1%) | 97 (3.7%) | 32 (12.7%) |
| Depression<sup>+</sup> | 253 (14.2%) | 40 (2.5%) | 23 (4.3%) | 140 (5.4%) | 23 (9.2%) |

<sup>\*</sup> Genotype data available, n(%): MESA 1716 (95.3%), ARIC 0 (0%), FHS-OS 388 (62.7%), MrOS 2296 (88.3%), SOF 139 (55.4%). <sup>+</sup> number (%). <sup>^</sup> mean (standard deviation). <sup>&</sup> median (interquartile range). <sup>§</sup> SOF participants with APOE genotyping came from only one study site. MESA – Multi-Ethnic Study of Atherosclerosis. ARIC – Atherosclerosis Risk in Communities. FHS-OS – Framingham Heart Study Offspring. MrOS – Osteoporotic Fractures in Men. SOF – Study of Osteoporotic Fractures. BMI – body mass index. APOE – apolipoprotein E. EEG – electroencephalography. BAI – Sleep EEG-based brain age index. AHI – Apnea-hypopnea index. IQR – interquartile range.

### BAI was positively associated with incident dementia

There were 1,082 incident dementia cases in total. For the minimally adjusted model, each 10-year increase in BAI was significantly associated with incident dementia at an HR of 1.39 (95% confidence interval [CI]: 1.20–1.60], p<0.001, I^2^ p = 0.33) (Figure 3A). After adjusting for age, sex, education, BMI, current smoking status, race, sleep medication, and physical activity levels, each 10-year increase in BAI was significantly associated with incident dementia at an HR of 1.39 (1.21-1.59, p<0.001, I^2^ p = 0.35) (Figure 3B). HRs across individual cohorts ranged from 1.19 (0.84-1.69, MESA) to 1.76 (1.07-2.90, SOF). Adding polynomials of BAI resulted in a better fit (lower Bayesian Information Criterion, BIC) in only one cohort (SOF), and no improvement in the other four cohorts.

**Figure 3.**
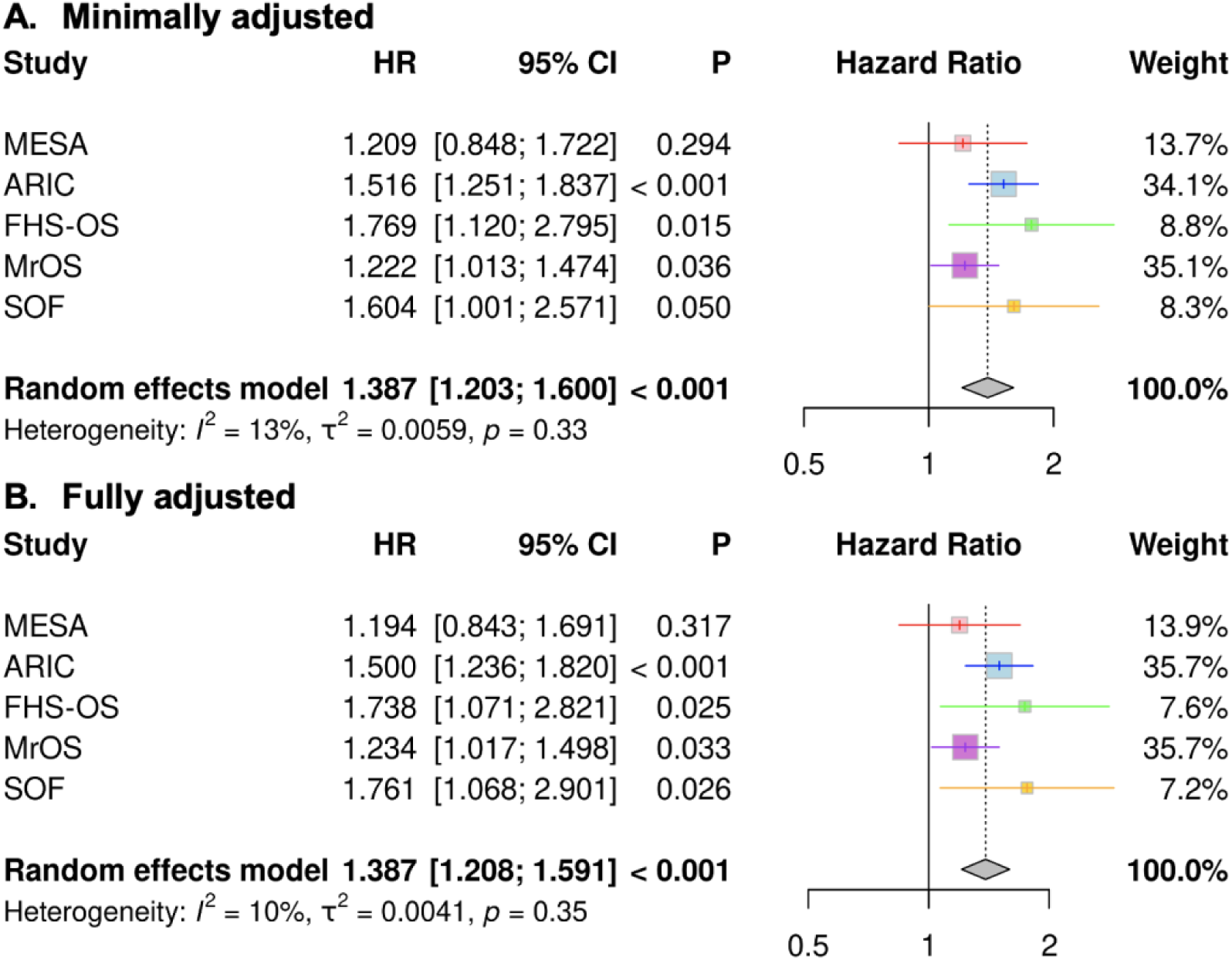
Pooled effect estimates for the association between brain age index and incident dementia. **(A)** adjusting for age and sex. **(B)** adjusting for age, sex, education, body mass index (BMI), current smoking status, race, sleep medication, and physical activity levels. CI – confidence interval. HR – hazards ratio. MESA – Multi-Ethnic Study of Atherosclerosis. ARIC – Atherosclerosis Risk in Communities. FHS-OS – Framingham Heart Study Offspring. MrOS – Osteoporotic Fractures in Men. SOF – Study of Osteoporotic Fractures.

Under the full adjustment model, we examined the individual sleep EEG microstructure features contributing to BAI (Figure 4). Ten out of thirteen features showed significant associations with incident dementia in the pooled analyses. Features from N2 and N3 showed a negative association with incident dementia, whereas the only N1 feature showed a positive association with incident dementia. The top feature in terms of statistical significance was waveform kurtosis in N2 (p<0.001), which was negatively associated with incident dementia and likely reflected K-complex activity, i.e., large, high-amplitude events that produce heavy-tailed amplitude distributions and are captured by high kurtosis.

**Figure 4.**
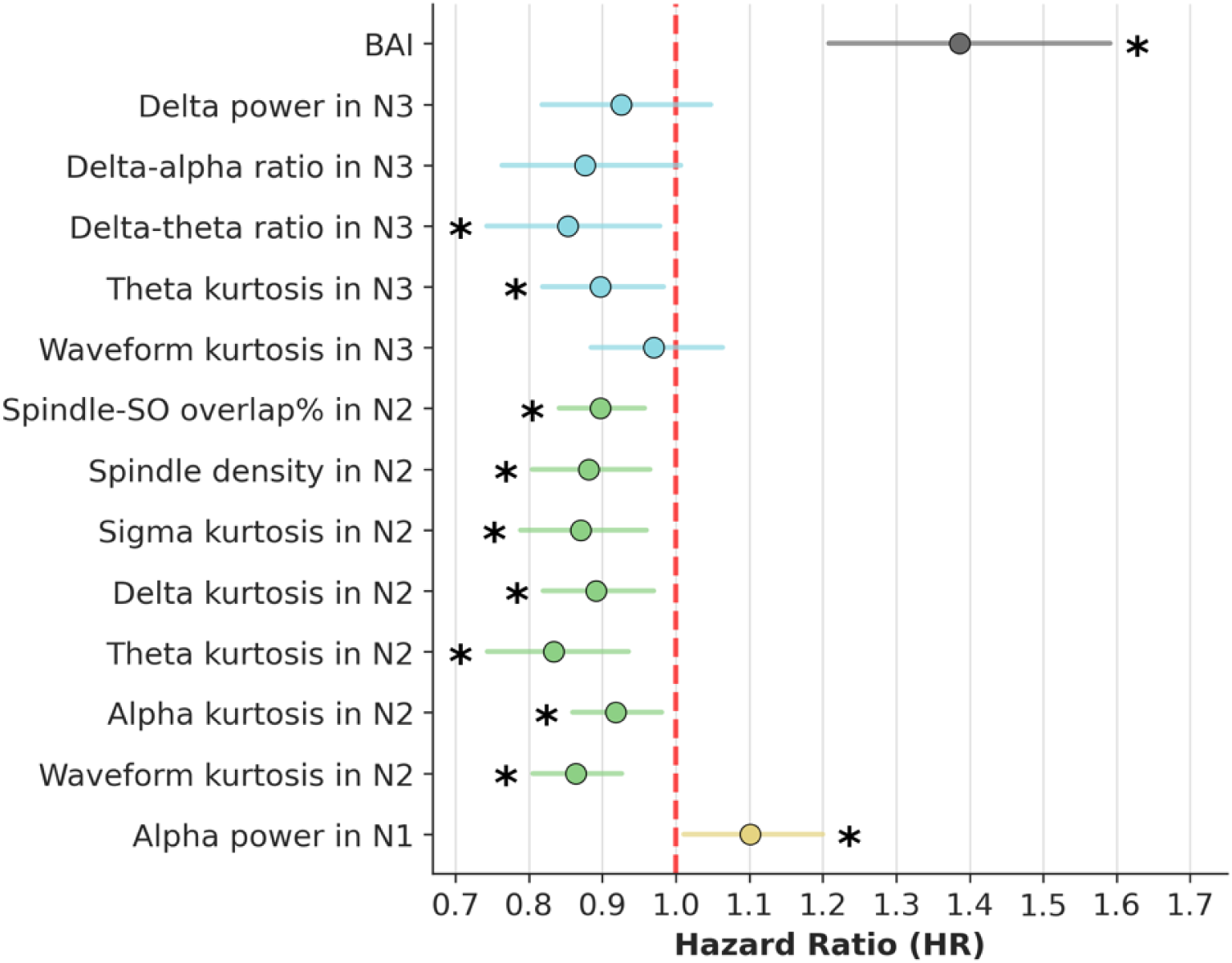
Pooled effect estimates for the associations between each individual EEG feature contributing to BAI and incident dementia. The x-axis is the hazard ratio (HR). HR>1 indicates a positive association with dementia risk; HR<1 indicates a negative association. BAI: 10-year increase. BAI features: 1-standard deviation increase.

### The BAI-dementia association remained in the sensitivity analyses

After further adjustment for diabetes, hypertension, heart attack, stroke, depression, baseline cognitive score, and AHI, the association between BAI and incident dementia was attenuated and remained significant (HR = 1.31 [1.14-1.50], p<0.001, Figure S1). Regarding APOE e4 allele, the association between BAI and dementia without adjusting for APOE e4 carrier was HR = 1.23 [1.04-1.46] (p=0.018), while the association remained significant after adjusting for APOE e4 carrier at HR = 1.22 [1.02-1.45] (p=0.027) (Figure S2A-B).

There was no interaction between age and BAI (interaction term HR = 1.06 [0.55-2.06], p=0.86), between sex and BAI (interaction term HR = 0.88 [0.49-1.57], p=0.66), or between APOE e4 carrier and BAI (interaction term HR = 0.94 [0.69-1.28], p=0.70). Stratified analyses showed that the HR was 1.65 in females (1.13-2.40, p=0.009, I^2^ p=0.03, N=2,447, N(incident dementia)=380, Figure S3A) and 1.25 in males (1.07-1.46, p=0.005, I^2^ p=0.89, N=4,624, N(incident dementia)=702, Figure S3B). Heterogeneity was observed only in the female-only analysis. Since the dementia outcome was identified using hospitalization information in MESA, with potential risk of underdiagnosis, we excluded MESA in sensitivity analyses. After excluding MESA, heterogeneity was no longer present, and the HR increased to 1.86 (95% CI: 1.52-2.28; p<0.001). The associations were similar among younger (<70 years old) (HR=1.43 [1.11-1.83], p=0.006, I^2^ p=0.31, N=3,288, N(incident dementia)=338; Figure S3C) and older individuals (≥70 years old) (HR=1.34 [1.12-1.60], p=0.001, I^2^ p=0.15, N=3,783, N(incident dementia)=744; Figure S3D).

## Discussion

Among 7,071 participants across five community-based cohorts, higher sleep EEG-based BAI was consistently associated with an increased risk of incident dementia over a median follow-up ranging from 3.5 to 17 years. Each 10-year increase in BAI was associated with a 39% increase in the risk of incident dementia. This association remained robust in various sensitivity analyses and stratified analyses. While a prior study has validated sleep-based BAI in clinical populations^21^, our findings show for the first time that the association between BAI and future dementia risk extends to community-dwelling populations.

The association between incident dementia and BAI based on sleep EEG microstructures (features) provides more granular information than the conventional sleep macrostructure measures. A recent pooled analysis^8^, including four of the five cohorts used in our study, found no significant associations between incident dementia and conventional sleep summary measures such as time spent in different sleep stages (pooled HRs for N1%, N2%, N3%, REM% ranged from 0.98 to 1.03, all p>0.05), wake after sleep onset, sleep efficiency, AHI, or relative delta power. These results suggested that traditional sleep macrostructure measures may not adequately capture the neurophysiological changes relevant to cognitive decline. This null result may reflect the somewhat arbitrary nature of several standard measures of sleep, including using 30-second scoring epochs and the criteria for slow waves to differentiate N2 vs. N3. Our study extends prior cross-sectional work in MESA and MrOS that compared sleep macrostructure and EEG microstructure in relation to cognitive performance^11^. While that study found EEG microstructures to be more strongly associated with cognition than macrostructure measures such as REM duration and sleep efficiency, the large number of EEG microstructure features limited clinical applicability. In contrast, we derived a single machine learning-based BAI that summarizes microstructural information and significantly predicts future dementia risk, advancing from cross-sectional associations to a longitudinal, potentially prognostic marker. However, the easy access of BAI depends on future works that integrate BAI into clinical systems and at-home settings^38,39^.

In our study, the association between BAI and dementia can be explained by several key EEG microstructures (features) that contribute to BAI, such as delta-to-theta ratio during N3, spindle density, and spindle-SO coupling during N2. Delta power, negatively associated with brain age^18^, is often linked to sleep quality^40^, synaptic homeostasis^41^, and glymphatic clearance^42^, all relevant for cognitive health. Spindle density, which is known to decline with age, reflects the integrity of thalamocortical circuits^19^. A reduction in spindle density in aging brains is associated with disrupted communication between the hippocampus and the cortex required for transferring memory into long-term storage^43^. Moreover, the loss of sleep spindles and spindle-SO coupling, both of which are linked to memory processing^44^, are associated with cognitive impairment and AD^45^, as captured by BAI. Meanwhile, multiple kurtosis-based EEG features showed significant negative associations with incident dementia, including theta kurtosis in N3, sigma/delta/theta/alpha kurtosis in N2, and waveform kurtosis in N2. Kurtosis is a measure of heavy-tailness, i.e., the extent of large outliers. One possible hypothesis is that they are related to cyclic alternating pattern (CAP), where the A1 phase consists of K-complexes and slow waves (in delta band) followed by spindles (in sigma band), the A3 phase consists of arousals (in alpha band), and the A2 phase (an intermediate phase between A1 and A3). Overall, the temporal organization of CAP and non-CAP may contribute to the kurtosis features underlying BAI, although this requires future verification. In addition, the theta kurtosis in N2 and N3 could be a result of theta bursts (TBs) preceding slow oscillations^46^, which have been associated with cognitive impairment and AD pathologies, including cerebrospinal fluid amyloid β 42 to 40 ratio and tau levels^47^. Meanwhile, alpha power during N1, a transitional state between wakefulness and sleep, is positively associated with incident dementia and brain age^20^. This increase in alpha activity reflects lighter sleep depth at this transitional state. On the other hand, there are EEG microstructures that are associated with dementia, but not being selected into the BAI model, which may be due to insufficient age dependence. For example, slow oscillation spatial involvement is associated with prodromal AD^48^. Many REM macrostructural and microstructural patterns are associated with cognitive impairment and/or AD pathology, such as REM latency^49^, theta power^50^, and theta-delta slowing^13^.

There are several limitations. First, the definitions of dementia and the length of follow-up varied across cohorts. For instance, in MESA, 86% of dementia outcomes were identified using ICD codes and death certificates from hospitalization, which may underreport true dementia cases; the remaining 14% were based on large declines in CASI scores (>1.5 SD decline) over six years, which does not reflect the actual time of dementia onset. In SOF, cognitive status was adjudicated only once, approximately 4-5 years after the PSG, where the time to diagnosis had to be approximated by time to adjudication, limiting precision in the dementia onset time. While such differences may affect comparability, we addressed this issue through random-effects meta-analysis, heterogeneity testing, and subgroup analyses. Second, only death was used as a competing risk for dementia. However, there are other conditions that may prevent the evaluation of dementia and/or be more likely to miss follow-up, such as surgeries. Third, due to the observational nature of these studies, we cannot infer a causal relationship between BAI and dementia. Randomized controlled trials are needed to evaluate whether interventions aimed at reducing BAI can slow cognitive decline.

## Conclusions

Across five large community cohorts, we found that an elevated sleep EEG-based BAI, a machine learning marker of brain aging, was independently linked to higher dementia risk. Using interpretable EEG microstructure features, BAI offers insights into neurophysiological signals that may influence future dementia risk or resilience.

## Acknowledgments

The authors thank all participants and staff of MESA, ARIC, FHS-OS, MrOS, and SOF cohorts for their essential contributions. This work is supported by NIH NIA R21AG085495 and R01AG083836 (PI Leng). Westover was supported by grants from the NIH (R01NS102190, R01NS102574, R01NS107291, RF1AG064312, RF1NS120947, R01AG073410), and NSF (2014431). Thomas was supported by an AASM Foundation Strategic Research Award and by the NIH (RF1AG064312, R01NS102190). Pase was supported by R01AG062531 as well as a National Health and Medical Research Council (NHMRC) Investigator Grant (GTN2009264).

The National Sleep Research Resource was supported by the National Heart, Lung, and Blood Institute (R24 HL114473, 75N92019R002).

MESA: The Multi-Ethnic Study of Atherosclerosis (MESA) Sleep Ancillary study was funded by NIH-NHLBI RO1 HL098433 and NIH-NIA 5R01AG070867. MESA is supported by NHLBI-funded contracts HHSN268201500003I, N01-HC-95159, N01-HC-95160, N01-HC-95161, N01-HC-95162, N01-HC-95163, N01-HC-95164, N01-HC-95165, N01-HC-95166, N01-HC-95167, N01-HC-95168, and N01-HC-95169 from the National Heart, Lung, and Blood Institute, and by cooperative agreements UL1-TR-000040, UL1-TR-001079, and UL1-TR-001420 funded by NCATS.

SHHS: The Sleep Heart Health Study (SHHS) was supported by National Heart, Lung, and Blood Institute cooperative agreements U01HL53916 (University of California, Davis), U01HL53931 (New York University), U01HL53934 (University of Minnesota), U01HL53937 and U01HL64360 (Johns Hopkins University), U01HL53938 (University of Arizona), U01HL53940 (University of Washington), U01HL53941 (Boston University), and U01HL63463 (Case Western Reserve University).

ARIC: ARIC was supported by National Heart, Lung, and Blood Institute cooperative agreements U01HL53934 (University of Minnesota) and U01HL64360 (Johns Hopkins University). The Atherosclerosis Risk in Communities study has been funded in whole or in part with Federal funds from the National Heart, Lung, and Blood Institute, National Institutes of Health, Department of Health and Human Services, under Contract nos (75N92022D00001, 75N92022D00002, 75N92022D00003, 75N92022D00004, 75N92022D00005). ARIC Neurocognitive (ARIC-NCS) for selected components of Visits 5 and 7 and all of Visits 6-8. The Atherosclerosis Risk in Communities Study is carried out as a collaborative study supported by National Heart, Lung, and Blood Institute contracts (75N92022D00001, 75N92022D00002, 75N92022D00003, 75N92022D00004, 75N92022D00005). The ARIC Neurocognitive Study is supported by U01HL096812, U01HL096814, U01HL096899, U01HL096902, and U01HL096917 from the NIH (NHLBI, NINDS, NIA, and NIDCD).

FHS: This work was supported by the National Heart, Lung, and Blood Institute contract (N01-HC-25195), National Institute on Aging (5R01-AG16495-03, AG059421, AG054076, AG049607, AG033090, AG066524, NS017950, P30AG066546, UF1NS125513), National Institute of Neurological Disorders and Stroke (5R01-NS17950-19 and 5R01-AG 08122), and the National Institutes of Health (N01-HC-25195, HHSN268201500001I, 75N92019D00031).

MrOS: The National Heart, Lung, and Blood Institute (NHLBI) provides funding for the MrOS Sleep ancillary study “Outcomes of Sleep Disorders in Older Men” under the following grant numbers: R01 HL071194, R01 HL070848, R01 HL070847, R01 HL070842, R01 HL070841, R01 HL070837, R01 HL070838, and R01 HL070839. The Osteoporotic Fractures in Men (MrOS) Study is supported by funding from the National Institutes of Health. The following institutes provide support: the National Institute on Aging (NIA), the National Institute of Arthritis and Musculoskeletal and Skin Diseases (NIAMS), and the National Center for Advancing Translational Sciences (NCATS) under the following grant numbers: U01 AG027810, U01 AG042124, U01 AG042139, U01 AG042140, U01 AG042143, U01 AG042145, U01 AG042168, U01 AR066160, R01 AG066671, and UL1 TR002369.

SOF: The Study of Osteoporotic Fractures (SOF) was supported by National Institutes of Health grants (AG021918, AG026720, AG05394, AG05407, AG08415, AR35582, AR35583, AR35584, RO1 AG005407, R01 AG027576-22, 2 R01 AG005394-22A1, 2 RO1 AG027574-22A1, HL40489, T32 AG000212-14).

## Disclosure

Westover is a co-founder, serves as a scientific advisor and consultant to, and has a personal equity interest in Beacon Biosignals. Thomas discloses: 1) patent and license/royalties from MyCardio, LLC, for the ECG-spectrogram; 2) patent and license/royalties from DeVilbiss-Drive for an auto-CPAP algorithm; 3) consulting for Jazz Pharmaceuticals, Guidepoint Global and GLG Councils. Other authors declare that they have no conflict of interest. SR has received consulting fees from Eli Lilly Inc., unrelated to this work, and is an unpaid scientific advisor to Apnimed.

## Data Availability

MESA can be requested from https://internal.mesa-nhlbi.org/. ARIC can be requested from https://biolincc.nhlbi.nih.gov/studies/aric/. FHS-OS can be requested from https://biolincc.nhlbi.nih.gov/studies/framoffspring/. The cognitive data of ARIC and FHS can be accessed via BioLINCC or dbGaP. MrOS can be requested from https://mrosonline.ucsf.edu/. SOF can be requested from https://sofonline.ucsf.edu/. The sleep signals for each cohort can be requested on the National Sleep Research Resource (NSRR): https://sleepdata.org.

## Code Availability

The code is available from GitHub: https://github.com/Hockey86/BAI-Dementia-Community-Cohorts.

## Supplemental Materials

### Supplementary Methods: Study Design

MESA is a multi-center, prospective cohort study of Black, White, Hispanic, and Chinese American adults. Between 2010 and 2012, participants were also enrolled in a Sleep Exam (in 2010-2013 in conjunction with MESA Exam 5), which included one full overnight unattended sleep polysomnography (PSG).

ARIC is a multi-center, prospective observational cohort study initially focused on cardiovascular risk factors, medical care, and disease in four US communities, between 1987 and 1989.

The FHS-OS cohort is a community-based prospective observational cohort study that enrolled offspring of the original FHS cohort from 1971 to 1975 in Framingham, Massachusetts^1^. Between 1995 and 1998, participants from the ARIC and FHS-OS cohorts completed overnight home-based polysomnography as part of the Sleep Heart Health Study (SHHS)^2^.

MrOS is a prospective study of community-dwelling individuals enrolled from 2000 to 2002 at six clinical centers in the United States. Sleep PSG data were collected between 2003 and 2005.

SOF is a multi-center prospective cohort study of women with sleep studies completed between 2002 and 2004.

### Supplementary Methods: Dementia Outcome Ascertainment

The primary outcome for our pooled analysis was incident dementia, with death treated as a competing risk. If neither dementia nor death occurred, the participant was censored at the last contact. The time-to-event was defined as the number of days from the sleep study to the first ascertainment of the event, including dementia, death, or other censoring events. Detailed definitions can be found in each cohort and are briefly described below.

In MESA^3^, the participants were followed up until 2018. Dementia was assessed using two criteria sequentially: (1) Possible International Classification of Diseases (ICD)-based all-cause dementia as adjudicated by the MESA Cognitive Working Group based on Fujiyoshi et al^4^; and (2) decrease in Cognitive Abilities Screening Instrument (CASI, a global cognitive score)^5^ from Exam 5 (close to sleep study) to Exam 6 is ≥ 1.5 standard deviations of the changes across participants, which was 9.8 points. The time to dementia is from the sleep study to the first date of hospitalization or death certificate, meeting any of the ICD-based criteria in case (1); and to Exam 6 in case (2). There is likely a substantial group of false negatives (i.e., outcome is falsely not dementia), from people who were never hospitalized or not known to be dead. Death was ascertained using death certificates from hospitals.

In ARIC^6^, the participants were followed up until 2019 (end of Visit 7). Dementia^7^ was based on reviewer diagnosis, algorithmic syndromic diagnosis, AD8 Dementia Screening Interview, Six Item Screener, dementia codes on the cohort eligibility form, and dementia codes on the death certificate form (ordered by priority, designated as level 3 in ARIC), while excluding discharge or death certificate codes that occurred after the date of last visit/phone contact. The time to dementia was based on the earliest date of the event and then subtracting 180 days to approximate the midpoint of the prior year when cognitive decline likely began. Death was ascertained through surveillance of death certificates from hospitals.

In FHS-OS^8^, the participants were followed up until 2018. Dementia^9^ was based on serial Mini-Mental State Examination (MMSE) from 1991-1998, and neuropsychological tests every 5 or 6 years since 1999. Participants who were identified as having possible dementia based on these screening assessments were invited to undergo additional, annual neurologic and neuropsychological examinations. Additional examinations were performed if the participant or a family member reported subjective cognitive decline. A dementia review panel reviewed every case of possible cognitive decline and dementia ever documented in the FHS, including the date of onset. The diagnosis of dementia was based on criteria from the Diagnostic and Statistical Manual of Mental Disorders (DSM-IV). All deaths were adjudicated by a panel, which reviews medical and nursing records up to the date of death and the death certificate.

In MrOS^10^, the participants were followed up until 2016 (end of Visit 4). Dementia was based on the report of physician-diagnosed dementia, self-reported dementia, dementia medication use, or having a change in the Modified Mini-Mental State (3MS) scores^11^ ≥ 1.5 standard deviations worse than the mean change from the sleep visit to any follow-up visit (a decline of 7.32, 9.43, and 13.62 points on 3MS from baseline to visit 2, visit 3 and visit 4, respectively). The 3MS is a global measurement of cognitive function, with components for orientation, concentration, language, praxis, and immediate and delayed memory. To ascertain death, participants were contacted every 4 months via postcard to determine vital status. Next of kin were contacted in cases of nonresponse. Reported deaths through 2018 were confirmed by a centralized review of death certificates. We used the death status at the end of Visit 4, the last visit at which cognitive status was obtained.

In SOF^12^, the dementia status was ascertained at Visit 9 (2007-2008). Dementia was identified through a two-stage protocol. First, all participants were screened for the following criteria: (1) score <88 on the 3MS; (2) score <4 on the CVLT delayed recall; (3) score ≥3.6 on the IQCODE; (4) previous dementia diagnosis; or (5) nursing home residence. These participants, along with a small random sample of screen-negative participants, underwent detailed adjudication by a rotating panel of dementia specialists who reviewed neuropsychological test results, functional measures, medication use, longitudinal cognitive data, mood scores, and medical history. The agreement among adjudicators achieved Cohen’s κ of 0.77. A diagnosis of dementia was made based on DSM-IV criteria, where the functional loss was based on IQCODE from the informant or, if unavailable, self-reported ADL/IADL limitations. Deaths were ascertained by contacts every 4 months and confirmed with death certificates. Four clinics individually collect SOF mortality, where a State Registered Certificate of Death was also submitted to the Coordinating Center.

---

**Figure S1.**
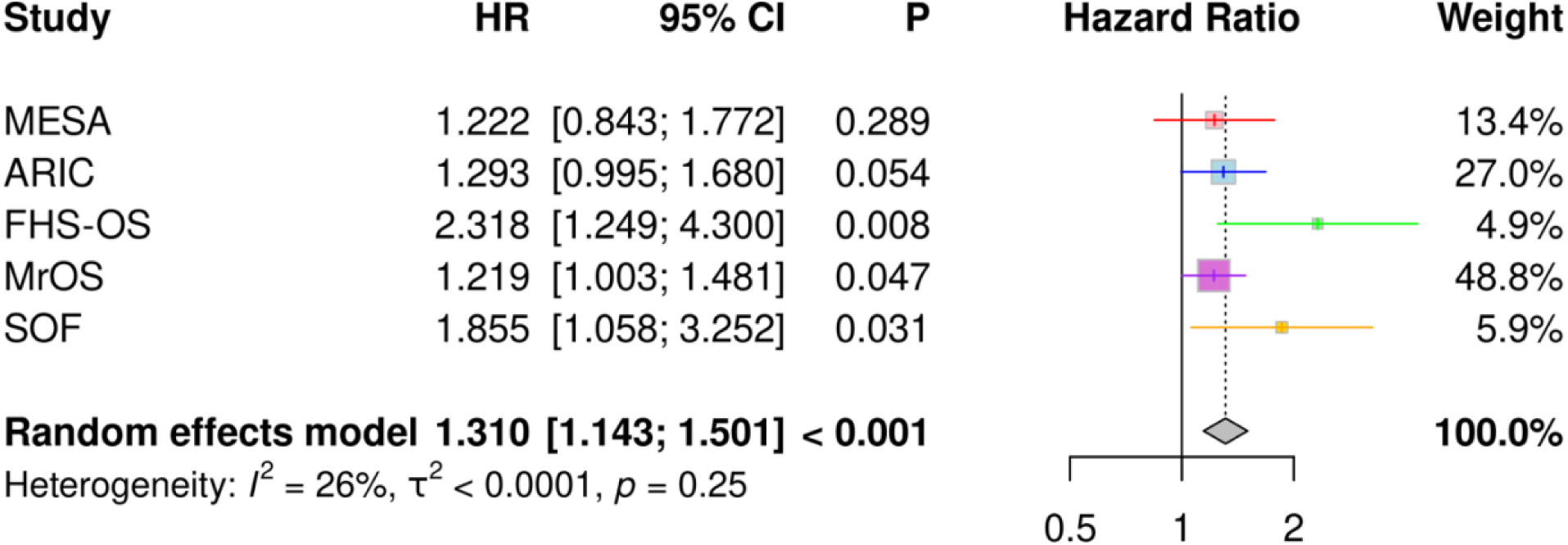
Forest plots of the associations between brain age index and incident dementia while adjusting for many (overcomplete) covariates, including age, sex, education, body mass index (BMI), current smoking status, race, sleep medication, physical exercise level, diabetes, hypertension, heart attack, stroke, depression, baseline cognitive score, and the apnea-hypopnea index, as a sensitivity analysis for the association between brain age index (BAI) and incident dementia. HR – hazards ratio. MESA – Multi-Ethnic Study of Atherosclerosis. ARIC – Atherosclerosis Risk in Communities. FHS-OS – Framingham Heart Study Offspring. MrOS – Osteoporotic Fractures in Men. SOF – Study of Osteoporotic Fractures.

**Figure S2.**
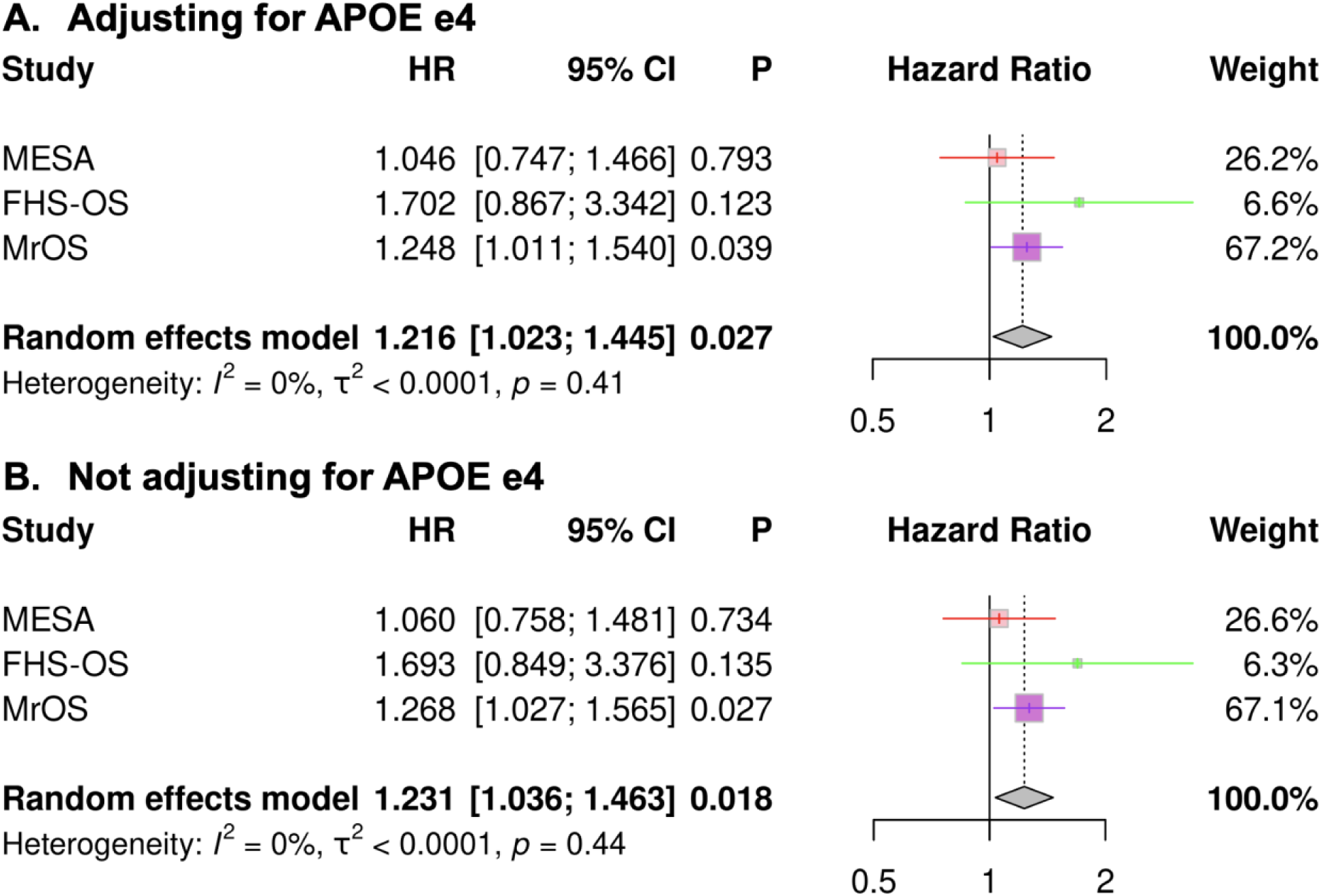
Forest plots of the associations between brain age index and dementia while adjusting (a) and not adjusting (b) APOE e4 allele carrier status to evaluate the relationship between brain age index (BAI) and incident dementia. For this analysis, the SOF cohort was excluded since the participants with APOE genotyping in SOF came from only one study site. Both analyses, with and without APOE, were performed based on the same subset of participants with APOE genotypes available. CI – confidence interval. HR – hazards ratio. MESA – Multi-Ethnic Study of Atherosclerosis. ARIC – Atherosclerosis Risk in Communities. FHS-OS – Framingham Heart Study Offspring. MrOS – Osteoporotic Fractures in Men. SOF – Study of Osteoporotic Fractures. * Only including the three cohorts with complete data on APOE e4 status.

**Figure S3.**
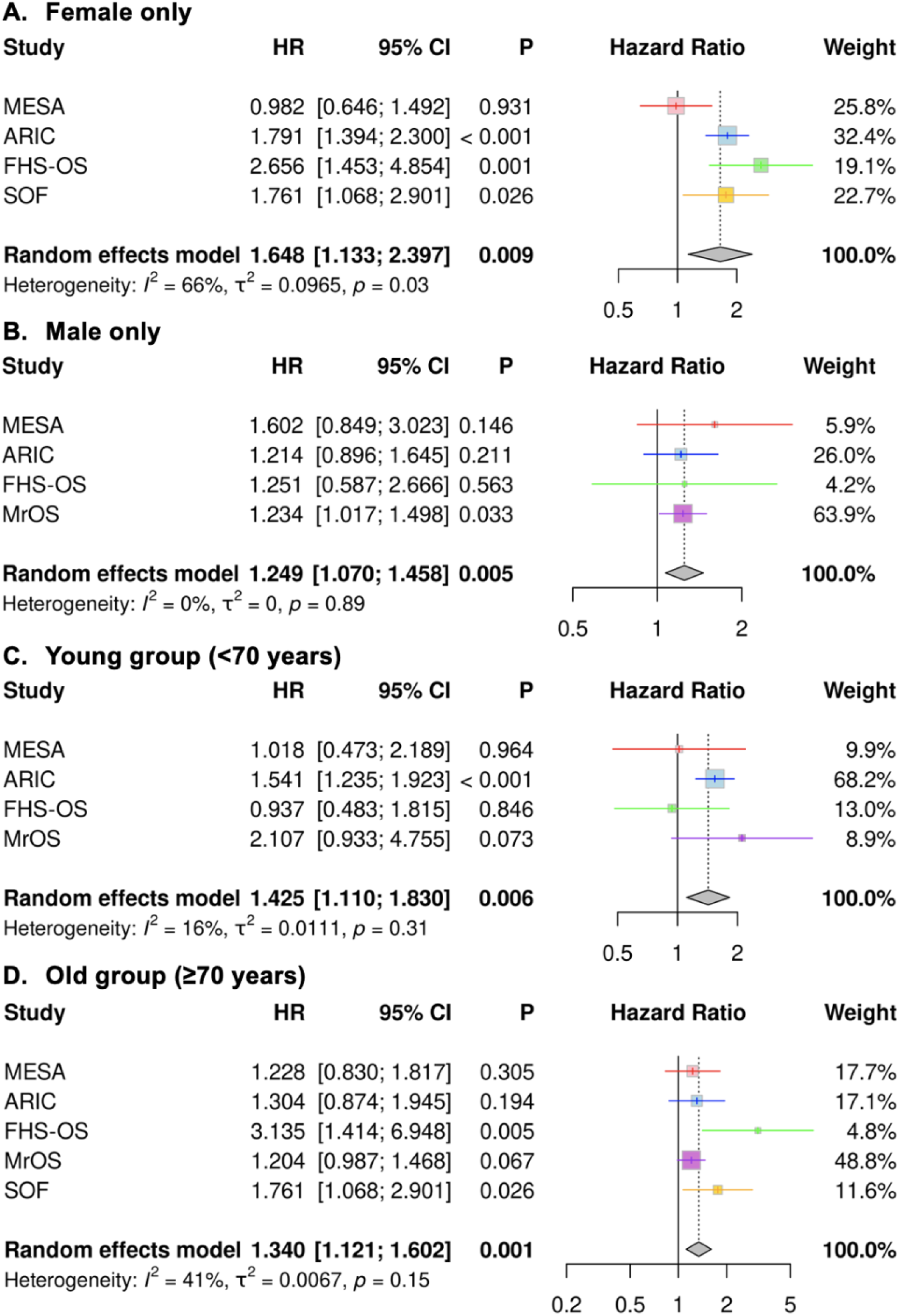
Forest plots of the associations between brain age index and incident dementia while stratified by sex (a-b) and age (c-d). Results were statistically significant across sex and age groups. SOF has all females. MrOS has all males. SOF has no participants under 70 years old. CI – confidence interval. HR – hazards ratio. MESA – Multi-Ethnic Study of Atherosclerosis. ARIC – Atherosclerosis Risk in Communities. FHS-OS – Framingham Heart Study Offspring. MrOS – Osteoporotic Fractures in Men. SOF – Study of Osteoporotic Fractures.

**Table S1.**
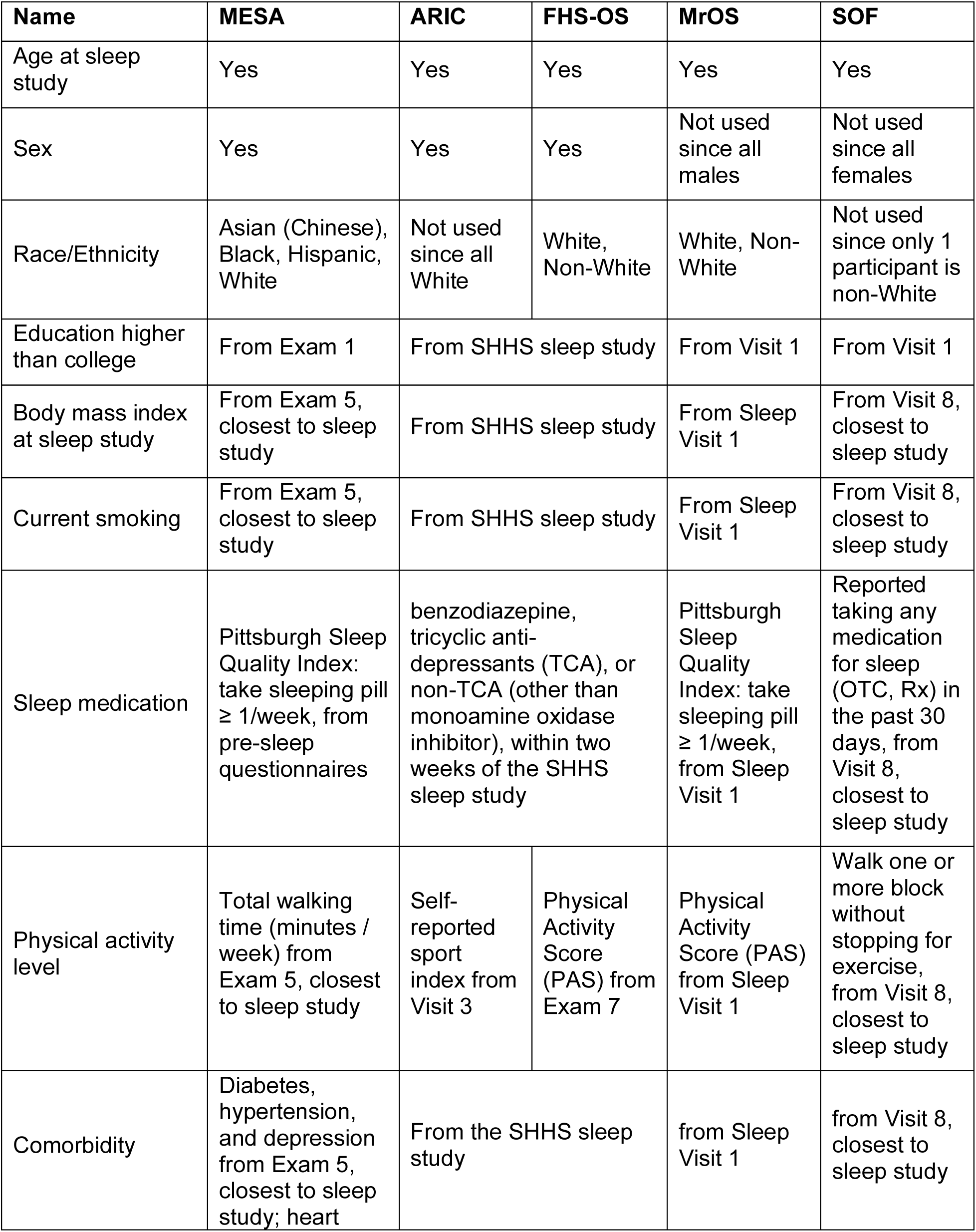
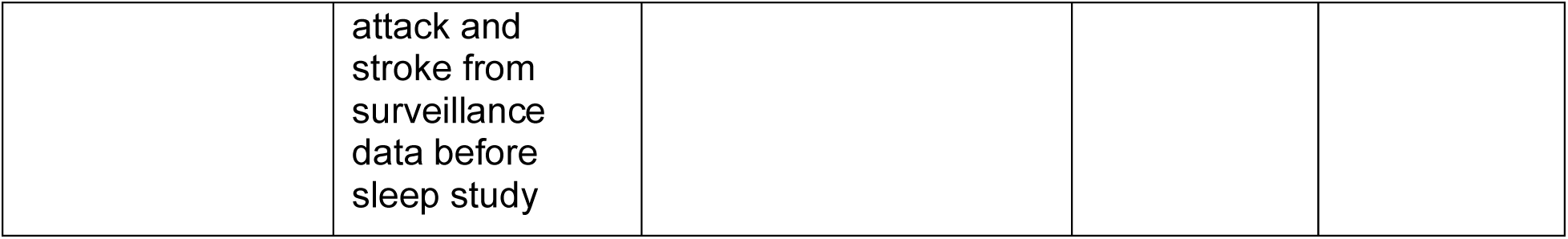
Covariates used in each cohort.

| Name | MESA | ARIC | FHS-OS | MrOS | SOF |
| --- | --- | --- | --- | --- | --- |
| Age at sleep study | Yes | Yes | Yes | Yes | Yes |
| Sex | Yes | Yes | Yes | Not used since all males | Not used since all females |
| Race/Ethnicity | Asian (Chinese), Black, Hispanic, White | Not used since all White | White, Non-White | White, Non-White | Not used since only 1 participant is non-White |
| Education higher than college | From Exam 1 | From SHHS sleep study |  | From Visit 1 | From Visit 1 |
| Body mass index at sleep study | From Exam 5, closest to sleep study | From SHHS sleep study |  | From Sleep Visit 1 | From Visit 8, closest to sleep study |
| Current smoking | From Exam 5, closest to sleep study | From SHHS sleep study |  | From Sleep Visit 1 | From Visit 8, closest to sleep study |
| Sleep medication | Pittsburgh Sleep Quality Index: take sleeping pill $\geq 1$/week, from pre-sleep questionnaires | benzodiazepine, tricyclic anti-depressants (TCA), or non-TCA (other than monoamine oxidase inhibitor), within two weeks of the SHHS sleep study | | Pittsburgh Sleep Quality Index: take sleeping pill $\geq 1$/week, from Sleep Visit 1 | Reported taking any medication for sleep (OTC, Rx) in the past 30 days, from Visit 8, closest to sleep study |
| Physical activity level | Total walking time (minutes / week) from Exam 5, closest to sleep study | Self-reported sport index from Visit 3 | Physical Activity Score (PAS) from Exam 7 | Physical Activity Score (PAS) from Sleep Visit 1 | Walk one or more block without stopping for exercise, from Visit 8, closest to sleep study |
| Comorbidity | Diabetes, hypertension, and depression from Exam 5, closest to sleep study; heart | From the SHHS sleep study |  | from Sleep Visit 1 | from Visit 8, closest to sleep study |

|  |  |
| --- | --- |
|  | attack and stroke from surveillance data before sleep study |

**Table S2.**
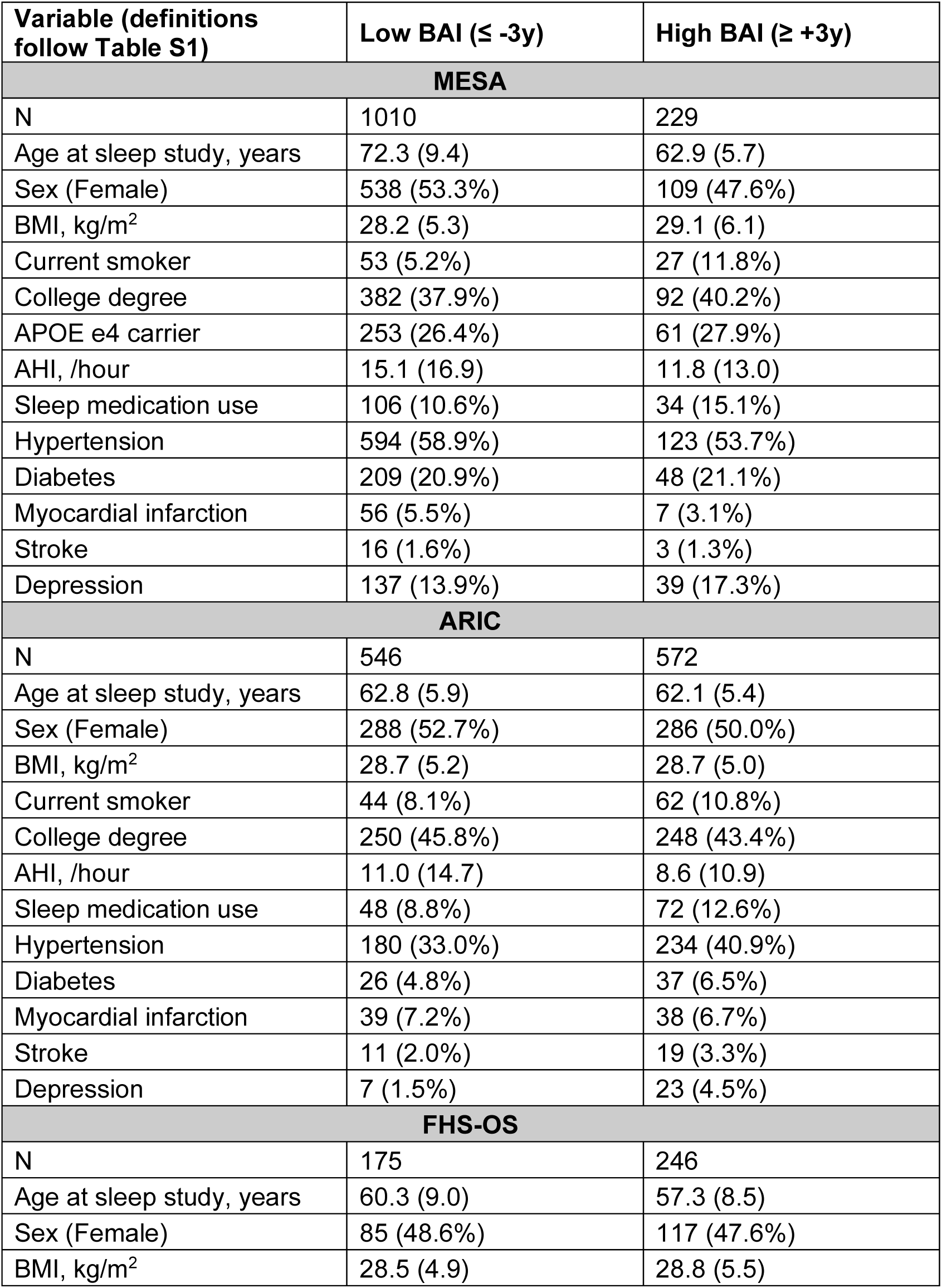
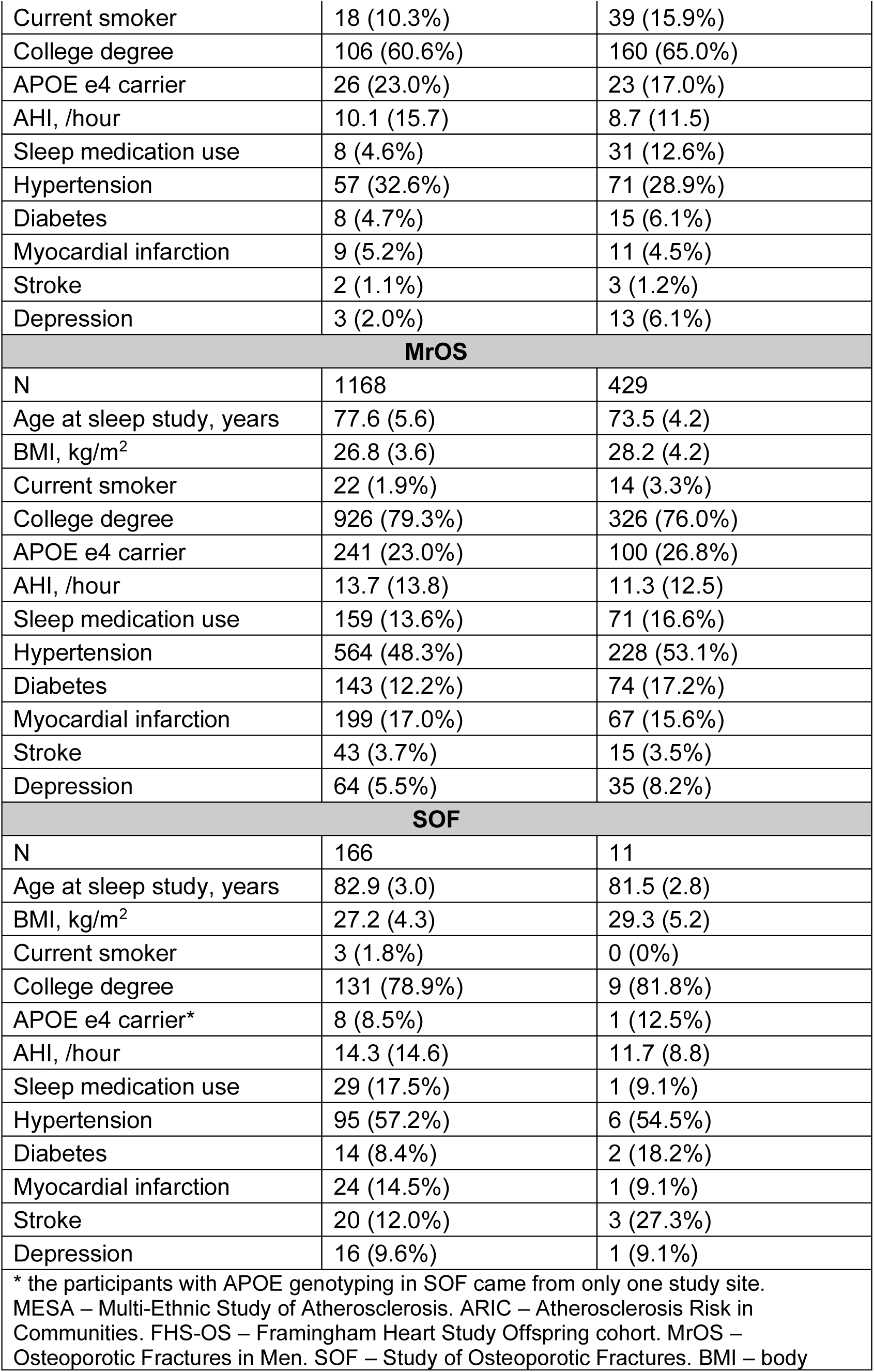
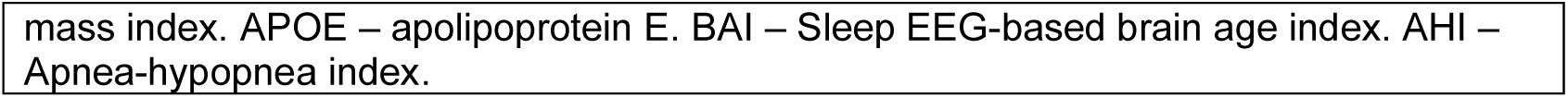
Participant characteristics for low BAI (≤ -3y) vs. high BAI (≥ +3y). In unadjusted analyses, participants with a lower BAI tended to be older across all cohorts (except SOF due to small sample size or the limited age range). No sex difference was observed. BMI was higher in the higher BAI group in MESA (28.2 vs. 29.1 kg/m², p=0.02) and MrOS (26.8 vs. 28.2 kg/m2, p<0.001). AHI was consistently lower in the higher BAI group in MESA, ARIC, and MrOS. Medications to improve sleep (sedative-hypnotics) were more commonly used in the higher BAI group in ARIC and FHS.

## Notes

### Author Declarations

MESA can be requested from https://internal.mesa-nhlbi.org/. ARIC can be requested from https://biolincc.nhlbi.nih.gov/studies/aric/. FHS-OS can be requested from https://biolincc.nhlbi.nih.gov/studies/framoffspring/. The cognitive data of ARIC and FHS can be accessed via BioLINCC or dbGaP. MrOS can be requested from https://mrosonline.ucsf.edu/. SOF can be requested from https://sofonline.ucsf.edu/. The sleep signals for each cohort can be requested on the National Sleep Research Resource (NSRR): https://sleepdata.org. These are individual-level data and are all de-identified.

